# Micronutrient deficiencies and the double burden of malnutrition in Vietnamese female adolescents: a national cross-sectional study in 2020

**DOI:** 10.1101/2024.04.11.24305546

**Authors:** Xiaomian Tan, Pui Yee Tan, Somphos Vicheth Som, Son Duy Nguyen, Do Thanh Tran, Nga Thuy Tran, Van Khanh Tran, J. Bernadette Moore, Yun Yun Gong

## Abstract

**Background:** Vietnam is facing a double burden of malnutrition, with increasing prevalence of overweight coexisting with undernutrition (stunting or thinness) and micronutrient deficiencies (MNDs). Although malnutrition during female adolescence leads to poor health outcomes with potential intergenerational effects on offspring, no studies have comprehensively investigated MNDs and nutritional status among contemporary Vietnamese female adolescents.

**Methods:** Data from 10-to 18-year-old female participants (n=1,471) in the nationally-representative Vietnam General Nutrition Survey 2020 were analysed. Blood nutritional biomarkers, anthropometric measurements, and socio-demographic data were collected and the associations between nutrition status and MNDs were analysed; with anaemia, iron deficiency (ID), iron deficiency anaemia, low serum zinc, low serum retinol, and any MNDs as specified outcomes.

**Findings:** Prevalence of overweight, stunting, and thinness was 27·2%, 14·3%, and 6·9%, respectively. Low serum zinc was common (39·8%), as was ID (9·8%). Bivariate analyses showed that older age (16-18 years old), ethnic minority, lower wealth index, and inflammation were associated with MNDs. In adjusted logistic regressions, stunting was associated with increased odds ratio (AOR) and [95% confidence intervals] of low serum retinol (8·92 [2·26, 35·15], p<0·01), as was thinness (12·25 [3·47, 43·33], p<0·01). Stunting was also associated with increased odds of having any MNDs (1·88 [1·13, 3·12], p<0·05).

**Interpretation:** More female adolescents were overweight than undernourished in Vietnam in 2020. However, undernutrition, low serum zinc, and ID remain prevalent. Food systems approaches should be considered to stem the stark increase in the double burden of malnutrition in young people living in Vietnam.

**Funding:** UK BBSRC BB/T008989/1.

## RESEARCH IN CONTEXT

### Evidence before this study

Malnutrition during female adolescence leads to poor health outcomes and has the potential for intergenerational effects on offspring. We searched PubMed from inception to February 1, 2024 for studies that had investigated micronutrient deficiencies (MNDs) or nutritional status (both over and undernutrition) in female adolescents living in Vietnam using the search terms “Vietnam*” and (“female*” or “girl*”) and “adolescent*” and (“double burden of malnutrition” or “micronutrient*” or “iron” or “zinc” or “vitamin A” or “overweight” or “obese” or “thinness” or “stunting” or “underweight”) with no language restriction. Of 188 studies identified, 104 had been published in last 10 years. These studies most frequently reported regional-specific data on body weight (City Ho Chi Minh or specific rural areas), as well as results from the Vietnamese cohorts in the Young Lives prospective cohort study which included children born in either 1994/5 or 2001/2 and were followed longitudinally for over 15 years since 2001. Although a few studies reported on the prevalence of stunting and iron deficiency in comparison with other Southeast Asian countries, we found no studies that had comprehensively investigated both micronutrient and nutritional status in contemporary Vietnamese adolescents.

### Added value of this study

This is the first study to report the prevalence of micronutrient deficiencies and the double burden of malnutrition in female adolecents living in Vietnam in 2020. Our study highlights the current double burden of malnutrition in Vietnam, with a higher prevalence of overweight (27·2%) than undernutrition (stunting and thinness combined at 21·2%) existing among contemporary female adolescents. In addition, our results show regional, residential, ethnic, and wealth index disparities in undernutrition and MNDs. Not least, our study suggests that iron deficiency and low serum zinc levels remain prevalent among Vietnamese female adolescents.

### Implications of all the available evidence

Additional efforts on targeted supplementation, food fortification, food-based interventions, and nutrition education should be made for at risk female adolescents living in Vietnam. Food environment and food systems approaches should be considered to stem the recent stark increase in overweight in young people living in Vietnam and other Western Pacific nations newly facing the double burden of malnutrition.

## INTRODUCTION

While considerable progress has been made in reducing the global burden of micronutrient deficiencies (MNDs) in recent decades, the prevalence of MNDs remains high in some populations.^1^ Concerningly, an estimated 56% of children under five years of age, and 69% of women of reproductive age (WRA, 15-49 years) globally are affected by at least one of a few, very common, MNDs.^2^ In particular, iron, zinc, and vitamin A deficiencies are the most common MNDs worldwide, and are causally associated with adverse health outcomes and the global burden of disease.^1,2^ Following infants and toddlers, female adolescents represent one of the most susceptible developmental and population age groups to MNDs.^3^

Indeed, after infancy and early childhood, adolescence is increasingly recognised as a unique and critical period for human growth and development, with profound hormonal, physical, and psychosocial changes.^4^ Poor nutrition during adolescence impacts full height development, quality of later life; and, in the case of adolescent females, has intergenerational effects on offspring that perpetuate an ongoing cycle of malnutrition.^3,4^ Approximately 1·8 billion individuals worldwide are adolescents between 10–19 years old, with 90% of them living in low-and middle-income countries (LMICs).^4^ In general, females have higher risk of MNDs such as iron deficiency (ID) and iron deficiency anaemia (IDA). This is because in addition to differences in reproductive biology, females are more susceptible to discriminative cultural, social, and gender norms.^3,4^ These discrimatory norms manifest in increased poverty, illiteracy, inequality, and early marriage, which individually and collectively increase risk for MNDs in female adolescents globally, and often particularly in LMICs.

Similar to other LMICs, Vietnam is facing a double burden of malnutrition, that is, the co-existence of under-and overnutrition at community and country levels.^5^ Economic development and rapid changes to food environment have resulted in a nutrition transition with a marked increase in the prevalence of overweight and obesity in childhood.^5,6^ Among 5–19-year-olds in Vietnam, the prevalence of overweight and obesity increased from 8·5% (2010) to 19% (2020).^7^ In Ho Chi Minh city, although the prevalence of stunting and thinness has decreased in recent decades, nonetheless undernutrition remained high at 6-14% for high school students in 2014; while 19% were overweight or obese.^5^ Similarly, although multiple national nutrition strategies aimed at improving population micronutrient status have been implemented,^8,9^ with consequent reductions in some MNDs, nonetheless the prevalence of some MNDs remains stubbornly high. For example, the pooled prevalence of anaemia in Vietnamese WRA decreased from 42·6% (1995) to 16·9% (2013), ^10^ but in 2010 zinc deficiency was noted to still be of severe public health significance, with more than 60% of WRA and children in Vietnam having low serum zinc.^11^

Given the rapid nutrition transition unfolding in Vietnam, the aims of this study were to examine: 1) the prevalence of different forms of malnutrition including undernutrition, overnutrition and MNDs (ID, low serum zinc and retinol); and 2) the associations between MNDs, demographic and socioeconomic factors, and growth indices; in female adolescents aged 10-18 years old, utilising the data from the nationally-representative, Vietnam General Nutrition Survey (GNS) 2020. Our hypothesis was that abnormal growth indices increase the risk of having MNDs.

## METHODS

### Study design

To understand the associations between MNDs and growth indices, GNS2020 data were analysed following the Strengthening the Reporting of Observational studies in Epidemiology (STROBE) guidelines. Conducted by the National Institute of Nutrition of Vietnam every 10 years, the GNS is a nationally-representative, population-based, cross-sectional survey that combines anthropometric and blood biomarker measurements with comprehensive sociodemographic data. In order to prevent selection bias, participants were recruited using a multi-stage cluster sampling design based on geographical areas, as detailed (currently in Preprint).^12^ Of the 20,864 participants completing the GNS 2020, a total of 1,471 participants met our inclusion criteria of being aged 10-to 18-years-old and female, were included in this study for analysis.

### Anthropometric measurements

Procedures for taking participant’s height and weight meaurements in the GNS 2020 have been described in detail in our Preprint.^12^ Body mass index (BMI) was calculated as weight (kg) divided by square of height (m^2^). The World Health Organization (WHO) growth reference was used to define stunting, thinness, and overweight.^13^ Stunting was defined by height-for-age z-score (HAZ)< -2 SD, while thinness and overweight were defined by BMI-for-age z score (BAZ)< -2 SD; and BAZ> +1, respectively. Outliers with HAZ > +6 SD or < -6 SD, or BAZ > +5 SD or < -5 SD were excluded from the dataset.

### Blood biomarkers and definition of micronutrient deficiencies

Blood samples were taken as part of the Vietnam GNS 2020 as detailed (Preprint)^12^. Biomarkers measured included: haemoglobin by point-of-care assay (HemoCue Diagnostics), serum ferritin and serum transferrin receptor by electrochemiluminescence immunoassay (Roche Cobas e 601; Roche Diagnostics), serum zinc by atomic emission spectrometry (MP-AES 4200, Agilent Technologies), serum retinol and retinol binding protein by high performance liquid chromatography (Agilent 1200, Agilent Technologies), serum c-reactive protein (CRP) and serum alpha-1-acid glycoprotein (AGP) using an automated protein analyzer (BN ProSpec Siemens® System, Siemens Healthineers). The diagnosis of MNDs followed the International Zinc Nutrition Consultative Group (IZiNCG)’s guidelines (for low serum zinc) and the WHO micronutrient survey manual (for other MNDs).^14,15^ Anaemia was defined as haemoglobin concentration levels <115 g/L and <120 g/L for 10-11 years old and 12-18 years old, respectively. For ID, the cut offs of serum ferritin concentration <15μg/L and 70μg/L were applied for apparently healthy individuals and individuals with infection or inflammation (CRP >5mg/L and/or AGP >1mg/L), respectively. IDA was defined as the co-existence of anaemia and ID. Low serum zinc was defined as serum zinc under different conditions: serum zinc <10·7μmol/L (morning, fasting), <10·1μmol/L (morning, non-fasting) or <9·0μmol/L (afternoon, non-fasting). Low serum retinol was defined as serum retinol concentration <0·70µmol/L. Any MNDs were defined as having at least one of any of the MNDs described above (ID, low serum zinc, and low serum retinol) and were reported as having 1 or ≥2 MNDs. To avoid bias in measurement, in addition to utilising the aforementioned standardised definitions and cut-offs from WHO and IZinNCG, within-assay and between-assay variability tests were performed for quality control of the biomarker measurements.

### Demographic and socioeconomic indicators

Demographic and socioeconomic data were collected using structured questionnaires. These included age, gender, geographical regions (Northern mountains, Delta and Hanoi, North Central and Central Coast, Highlands, Southeast and City Ho Chi Minh, and Mekong Delta), urban or rural demographics, ethnicity (Kinh or a minority ethinic group, e.g. Tay, Thai, Muong, Khmer, Nung, Hmong or other), and household wealth index (quintiles). The details of the wealth index calculation have been reported (preprint).^12^

### Statistical analyses

Descriptive and logistic regression analyses were performed using STATA 17 (Stata Corporation, College Station, TX, US). All analyses were performed based on sampling weights, using the svy prefix. The number and percentage of missed observations within each variable are listed in **Table S1**. Quantitative data are presented as mean ± SD for continuous variables and number (%) for categorical variables, with the exception of serum ferritin levels, which are presented as geometric mean. Given the large sample size, the Wald and Chi-Square tests were performed on continuous and categorical variables, respectively, to assess the difference of characteristics of participants among nutritional status, unless specific noted. Bivariate logistic regression was performed to assess the associations between MNDs (outcomes) and growth indices and demographic and socioeconomic indicators. Multivariate logistic regression was used to investigate the associations between micronutrient status and growth indices, after adjusting for confounders: age group (done categorically for groups 10-12, 13-15, 16-18 years old), inflammation and sociodemographic factors (area of residence, ethnicity and wealth index). Crude (COR) and adjusted odds ratio (AOR) with 95% confidence intervals (CI) were reported for binominal outcomes (5 individual MNDs), while crude (CRR) and adjusted relative risk (ARR) with 95% CI were reported for multi-nominal outcomes (any 1 or more of any MNDs). Significant level was set to 0·05. Sensitivity analyses were done by: 1) comparing subsamples (50% of the total population, randomly picked) with the total sample; and 2) testing 2 different logistic regression models with or without inflammation status as a covariate. Data visualisation was conducted using ggplot2 in the R environment (R-4·2·3).

## RESULTS

### Characteristics of participants

Data from 1,471 female adolescent participants in the Vietnam GNS 2020, aged 10-18 years old were included in this study. The overall prevalence of overweight, stunting, and thinness was 27·2%, 14·3%, and 6·9%, respectively (**Table 1**). Sociodemographic factors investigated in relation to nutritional status included geographical areas, urban or rural areas, ethnicity, and wealth index. These were all associated with stunting (p<0·01, **Table 1**). Stunting was more prevalent in: adolescents living in northern mountains, north central & central coast and highlands; rural areas; ethnic minorities; and those living in households with a low wealth index. In contrast, high prevalence of overweight (24·4%) and thinness (22·4%) was found among females who lived in Delta and Hanoi. Prevalence of overweight was low in Northern mountains (8·6%) while thinness prevalence (15·3%) was high in this area (**Table 1**). The average age of the study participants was 12·8 ± 2·2 years old, although adolescents with stunting were older than those without stunting (13·3 ± 2·6 vs 12·6 ± 2·1, p<0·01). While thin adolescents were younger than those with normal weight (12·1 ± 2·0 vs 13·0 ± 2·2, p< 0·01).

**Table 1.** Socio-demographic characteristics of female adolescents living in Vietnam between 10 to 18 years old (n=1471).

|  | Total<br>(n=1471) | Non-stunted<br>(n=1068; 85.7%) | Stunted<br>(n=185; 14.3%) | P value | NW<br>(n=967; 65.9%) | OW<br>(n=402; 27.2%) | Thinness<br>(n=102; 6.9%) | P value |
| --- | --- | --- | --- | --- | --- | --- | --- | --- |
| Age in years, mean ± SD | 12.8 ± 2.2 | 12.6 ± 2.1 | 13.3 ± 2.6 | <0.01<sup>a</sup> | 13.0 ± 2.2 | 12.6 ± 2.2 | 12.1 ± 2.0 | <0.01<sup>b</sup> |
| Geographical region, n (%) |  |  |  |  |  |  |  |  |
| Northern mountains | 208 (14.5%) | 132 (13.1%) | 57 (30.4%) | <0.01 | 159 (16.9%) | 38 (8.6%) | 11 (15.3%) | 0.51 |
| Delta and Hanoi | 306 (26.1%) | 265 (28.5%) | 19 (14.1%) |  | 233 (27.1%) | 56 (24.4%) | 17 (22.4%) |  |
| North Central & Central Coast | 262 (19.1%) | 188 (18.2%) | 40 (26.6%) |  | 174 (19.6%) | 69 (17.6%) | 19 (19.8%) |  |
| Highlands | 271 (8.0%) | 151 (6%) | 34 (9%) |  | 140 (6.3%) | 110 (11.9%) | 21 (9.4%) |  |
| Southeast and City Ho Chi Minh | 220 (16.2%) | 168 (15.8%) | 16 (10.4%) |  | 138 (14.9%) | 74 (21.4%) | 8 (8.2%) |  |
| Mekong Delta | 204 (16.1%) | 164 (18.4%) | 19 (9.5%) |  | 123 (15.1%) | 55 (16.1%) | 26 (25%) |  |
| Area of residence, n (%) |  |  |  |  |  |  |  |  |
| Urban | 614 (35.7%) | 454 (36.2%) | 48 (19.5%) | <0.01 | 377 (32.3%) | 199 (44.2%) | 38 (34.1%) | 0.10 |
| Rural | 857 (64.3%) | 614 (63.8%) | 137 (80.5%) |  | 590 (67.7%) | 203 (55.8%) | 64 (65.9%) |  |
| Ethnicity, n (%) |  |  |  |  |  |  |  |  |
| Kinh | 1171 (82.2%) | 907 (87.7%) | 96 (58.8%) | <0.01 | 768 (82.9%) | 325 (80.6%) | 78 (81.6%) | 0.82 |
| Minorities | 300 (17.8%) | 161 (12.3%) | 89 (41.2%) |  | 199 (17.1%) | 77 (19.4%) | 24 (18.4%) |  |
| Wealth index, n (%) |  |  |  |  |  |  |  |  |
| Lowest | 247 (17.5%) | 149 (15.5%) | 67 (31.8%) | <0.01 | 177 (18.6%) | 53 (14.6%) | 17 (18%) | 0.71 |
| Second | 304 (17.4%) | 211 (17%) | 48 (22.2%) |  | 187 (17%) | 87 (17.2%) | 30 (22.1%) |  |
| Third | 323 (22.2%) | 228 (21.7%) | 34 (23.9%) |  | 198 (21.3%) | 100 (22.9%) | 25 (27.9%) |  |
| Fourth | 351 (25.2%) | 273 (26.3%) | 22 (14.5%) |  | 227 (24.6%) | 109 (27.8%) | 15 (20.2%) |  |
| Highest | 246 (17.8%) | 207 (19.4%) | 14 (7.7%) |  | 178 (18.5%) | 53 (17.5%) | 15 (11.8%) |  |
| Data are presented as mean ± SD for continuous variables and n (%) for categorical variables.<br>Wald test and Chi square test were performed for continuous variables (age in year) and categorical variables (Geographical region, area of residence, ethnicity and wealth index), respectively.<br><sup>a</sup>P value was calculated comparing with non-stunted group using Wald test; <sup>b</sup>P value calculated comparing thinness group with NW group, by Wald test.<br>NW: normal weight; OW: overweight. | | | | | | | | |

Interestingly, no differences were observed in the levels of serum biomarkers between stunted and non-stunted adolescents (**Table 2**). However, both serum retinol and retinol binding protein levels were significantly lower in adolescents who were thin compared to those with normal weight (p<0·01, **Table 2**). While 5·9% of participants exhibited markers of either acute or chronic inflammation, these indicators did not vary significantly in relation to nutritional status. No differences were observed in blood biomarkers between the overweight and normal weight groups. Notably, serum ferritin levels were higher in participants with thinness (p<0·01), but remarkably ID was not associated with body weight status (**Table 2**). Prevalence of low serum retinol was much higher in adolescents with thinness compared to those with overweight and normal weight (7·0% versus 1·7% and 0·8%; p<0·01); and was found more prevalent among adolescents with stunting than those who were not stunted (3·8% vesus 1·0%, p=0·04). Moreover, having any individual or combination of MNDs were associated with stunting (p<0·01) but not thinness or overweight (**Table 2**).

**Table 2.** Anthropometric, biochemical characteristics and micronutrient status of female adolescents living in Vietnam between 10 to 18 years old (n=1471).

|  | Total<br>(n=1471) | Non-stunted<br>(n=1068; 85·7%) | Stunted<br>(n=185; 14·3%) | P value<sup>a</sup> | NW<br>(n=967; 65·9%) | OW<br>(n=402;<br>27·2%) | P value<sup>b</sup> | Thinness<br>(n=102; 6·9%) | P value<sup>b</sup> |
| --- | --- | --- | --- | --- | --- | --- | --- | --- | --- |
| Anthropometric |  |  |  |  |  |  |  |  |  |
| HAZ | -0·94 ± 1·07 | -0·65 ± 0·82 | -2·67 ± 0·62 | <0·01 | -0·99 ± 1 | -0·46 ± 1·2 | <0·01 | -1·38 ± 1·07 | 0·02 |
| BAZ | -0·31 ± 1·22 | -0·23 ± 1·2 | -0·78 ± 1·21 | <0·01 | -0·42 ± 0·79 | 1·56 ± 0·46 | <0·01 | -2·61 ± 0·52 | <0·01 |
| Biomarkers |  |  |  |  |  |  |  |  |  |
| Haemoglobin (g/dL) | 13·06 ± 1·03 | 13·10 ± 0·97 | 12·79 ± 1·25 | 0·09 | 13·06 ± 1·04 | 13·12 ± 0·98 | 0·39 | 12·9 ± 1 | 0·13 |
| Serum ferritin (µg/L)<br>(geometric mean ± SE)<sup>c</sup> | 54·76 ± 1·16 | 56·78 ± 1·58 | 52·76 ± 5·45 | 0·51 | 53·69 ± 1·90 | 54·43 ± 2·43 | 0·85 | 67·47 ± 4·41 | <0·01 |
| Serum transferrin receptor<br>(mg/L) | 5·87 ± 2·55 | 5·71 ± 1·85 | 6·55 ± 4·61 | 0·13 | 5·87 ± 2·55 | 5·94 ± 2·6 | 0·77 | 5·64 ± 2·32 | 0·26 |
| Serum zinc (µmol/L) | 11·37 ± 5·32 | 11·39 ± 5·22 | 11·3 ± 6·21 | 0·89 | 11·35 ± 5·5 | 11·51 ± 5·07 | 0·72 | 11·12 ± 4·39 | 0·74 |
| Serum retinol (µmol/L) | 1·25 ± 0·29 | 1·25 ± 0·29 | 1·22 ± 0·31 | 0·43 | 1·25 ± 0·29 | 1·27 ± 0·3 | 0·58 | 1·14 ± 0·28 | <0·01 |
| RBP (µmol/L) | 1·37 ± 0·44 | 1·37 ± 0·45 | 1·35 ± 0·39 | 0·55 | 1·38 ± 0·45 | 1·39 ± 0·44 | 0·77 | 1·18 ± 0·3 | <0·01 |
| CRP (mg/L) | 0·62 ± 2·79 | 0·62 ± 2·54 | 0·85 ± 4·9 | 0·56 | 0·49 ± 2·6 | 0·86 ± 2·65 | 0·12 | 0·94 ± 4·51 | 0·46 |
| AGP (mg/L) | 0·56 ± 0·25 | 0·56 ± 0·24 | 0·59 ± 0·32 | 0·34 | 0·55 ± 0·24 | 0·59 ± 0·26 | 0·13 | 0·51 ± 0·25 | 0·23 |
| MNDs |  |  |  |  |  |  |  |  |  |
|  | Total<br>(n=1471) | Non-stunted<br>(n=1068; 85·7%) | Stunted<br>(n=185; 14·3%) | P value | NW<br>(n=967; 65·9%) | OW<br>(n=402; 27·2%) |  | Thinness<br>(n=102; 6·9%) | P value |
| Anaemia | 143 (9·5%) | 94 (8·8%) | 29 (14·8%) | 0·07 | 97 (10%) | 34 (7·1%) |  | 12 (13·8%) | 0·14 |
| ID | 136 (9·8%) | 93 (8·6%) | 17 (9·9%) | 0·66 | 94 (10%) | 39 (11·3%) |  | 3 (2·1%) | 0·17 |
| IDA | 62 (4·2%) | 39 (3·5%) | 12 (6·3%) | 0·07 | 43 (4·5%) | 18 (4·4%) |  | 1 (1·1%) | 0·43 |
| Low serum zinc | 563 (39·8%) | 397 (39·5%) | 83 (49·6%) | 0·11 | 375 (41·5%) | 151 (35·1%) |  | 37 (42·5%) | 0·21 |
| Low serum retinol | 16 (1·5%) | 9 (1·0%) | 3 (3·8%) | 0·04 | 7 (0·8%) | 6 (1·7%) |  | 3 (7·0%) | <0·01 |
| Chronic or acute inflammation | 82 (5·9%) | 57 (5·3%) | 12 (6·8%) | 0·51 | 49 (4·6%) | 28 (9%) |  | 5 (5%) | 0·09 |
| Number of any MNDs<sup>d</sup> |  |  |  |  |  |  |  |  |  |
| 0 | 412 (53·9%) | 369 (56·3%) | 43 (38·3%) | <0·01 | 306 (52·3%) | 135 (60·7%) |  | 33 (48·7%) | 0·37 |
| 1 | 306 (41·6%) | 262 (39·8%) | 44 (53·3%) |  | 239 (43·1%) | 92 (34·0%) |  | 23 (47·6%) |  |
| 2 | 30 (4·4%) | 22 (3·8%) | 8 (8·4%) |  | 24 (4·5%) | 11 (4·6%) |  | 2 (3·7%) |  |
| 3 | 1 (0·1%) | 1 (0·1%) | 0 (0%) |  | 1 (0·1%) | 2 (0·7%) |  | 0 (0%) |  |
| Data are presented as mean ± SD for continuous variables and n (%) for categorical variables unless specific noted.<br>Wald test and Chi square test were performed for continuous variables (anthropometric, biomarkers) and categorical variables (MNDs and number of any MNDs), respectively.<br><sup>a</sup>P value was calculated comparing with non-stunted group; <sup>b</sup>P value was calculated comparing with normal weight group; <sup>c</sup>P value was calculated based on a univariate regression model; <sup>d</sup>Participants with numbers of any following MNDs: ID, Low serum zinc and retinol.<br>AGP: alpha-1 acid glycoprotein; BAZ: BMI-for-age z score; CRP: C-reactive protein; HAZ: height-for-age z score; NW: normal weight; OW: overweight; RBP: retinol binding protein. |  |  |  |  |  |  |  |  |  |

### Prevalence of malnutrition and micronutrient deficiencies

Notable differences in nutritional status and MNDs emerged when participants were stratified into early-adolescent (10-12 years old), middle-adolescent (13-15 years old) and late-adolescent (16-18 years old) age groups (**Figure 2**). Prevalence of both overweight and thinness was higher in early-adolescents compared to late adolescents (29·7% versus 22·0%, 9·4% versus 2·7%, respectively; **Figure 2A**). Whereas, compared to early-adolescents, the prevalence of stunting was higher in the late adolescent group (26·8% versus 12·4%; **Figure 2A**). Anaemia prevalence was lowest in the early-adolescent group (6·8%), but more common in late adolescents (14·1%; **Figure 2B**). Similarly, the prevalence of IDA was dramatically higher in middle-(5·6%) and late-(9·0%) adolescents in comparison to early-adolescent females (2·1%; **Figure 2B**). Both ID and low serum zinc were more prevalent in the late adolescents, whereas low serum retinol was less prevalent in the oldest group (**Figure 2C**). Low serum zinc was the most common MND, affecting 39·8% of the study participants, particularly in late adolescents (47·7%; **Figure 2C**). Only 1·5% of the female adolescents had low serum retinol levels (**Figure 2C**). When we examined the existence of any MNDs, nearly half (45·6%) of all female adolescents in the GNS 2020 had at least one MND (**Figure 2D**). The prevalence of having ≥1 MNDs was lowest in early adolescents (43·8%) and highest in late adolescents (55·6%; **Figure 2D**). Notably late adolescents had a much higher prevalence of having 2 MNDs (11·3%) compared to early adolescents (3·0%). Only a very small proportion (0·3%) of study participants had three MNDs (**Figure 2D**).

**Figure 1.**
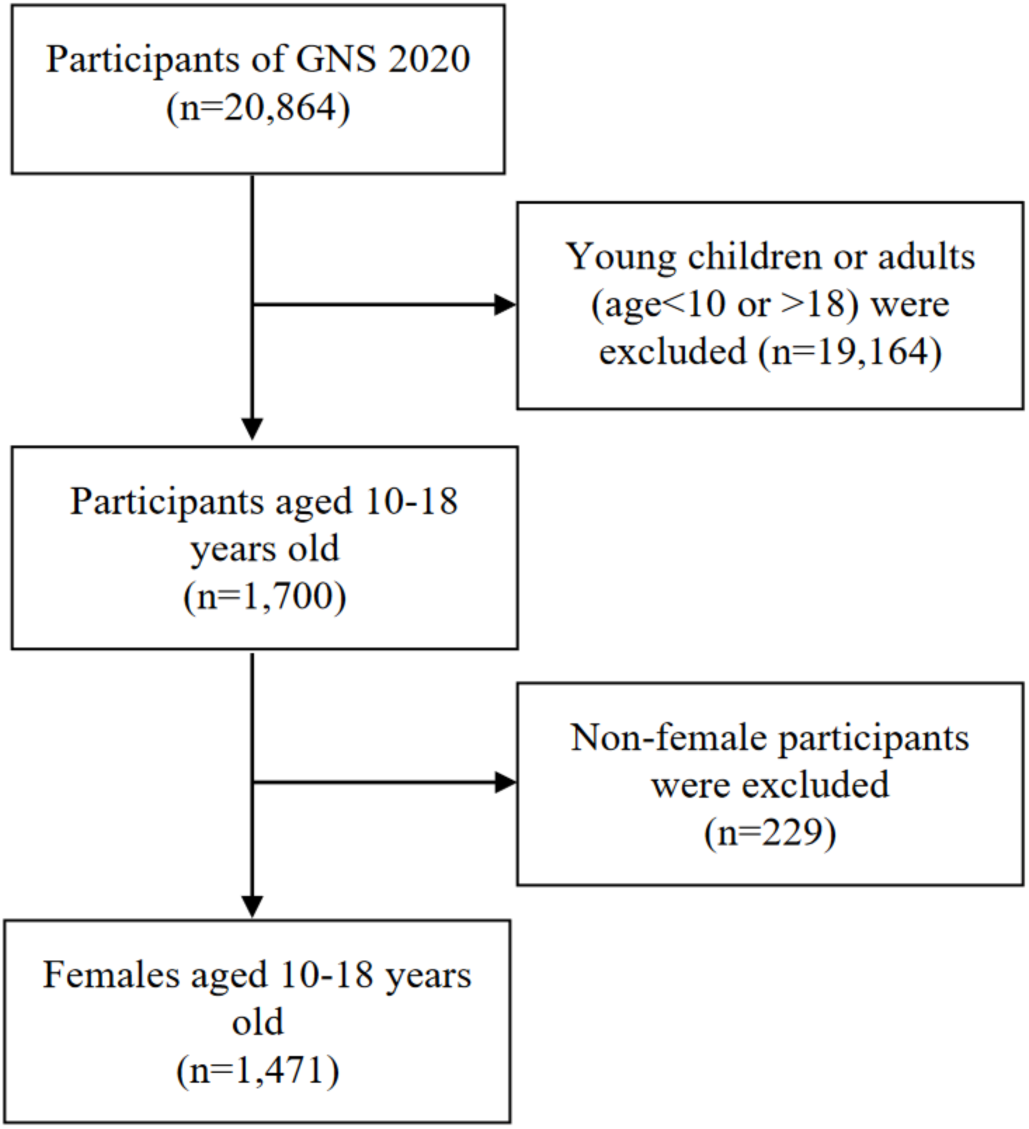
Participants selection flow chart.

**Figure 2.**
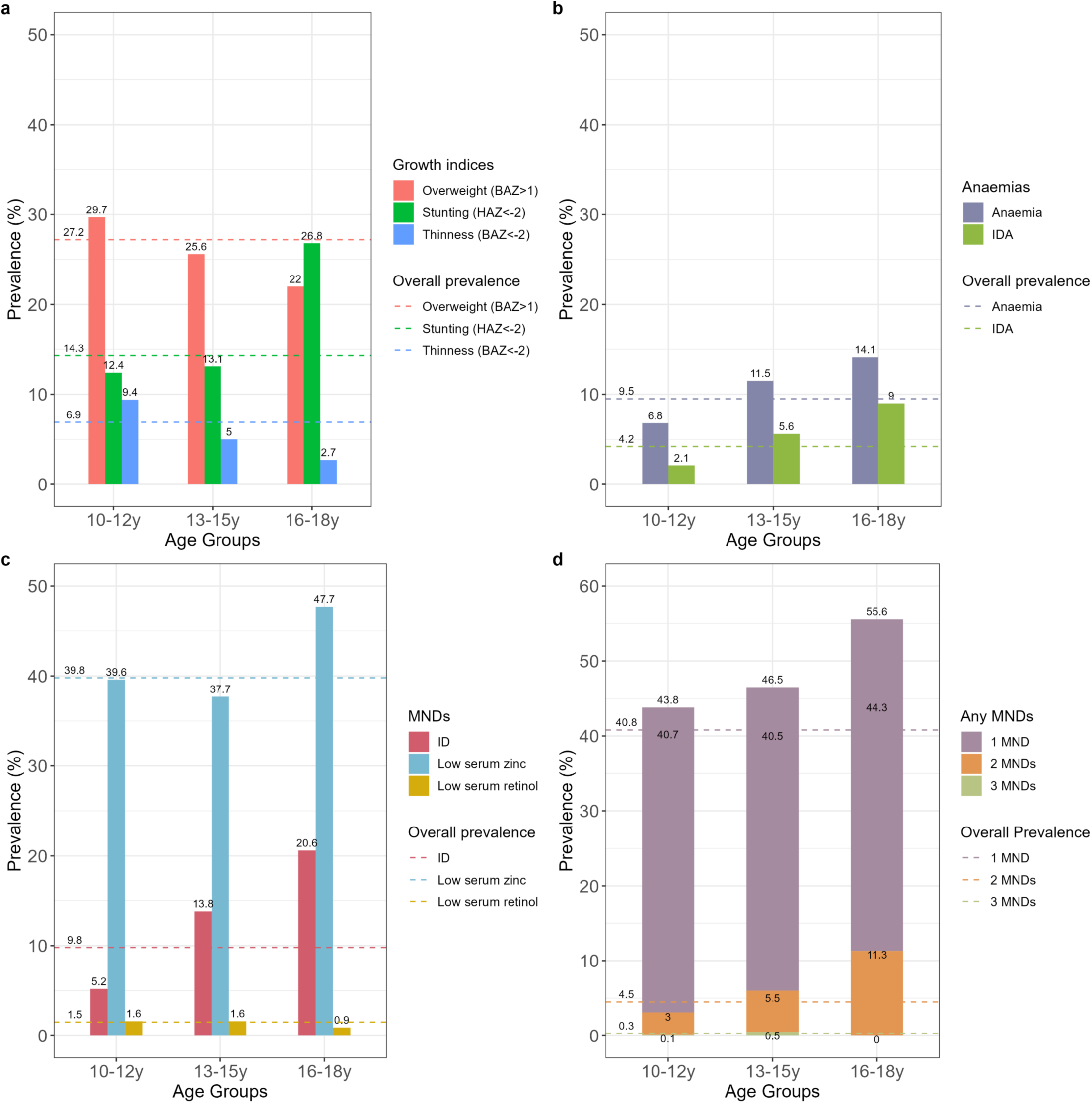
Prevalence of malnutrition in female adolescents aged 10-18 years old living in Vietnam in different age groups. Data show the prevalence of overweight, stunting and thinness **(a)**, anaemias **(b)**, MNDs **(c)** and distribution of any MNDs (any of ID, low serum zinc or low serum retinol) **(d)**. Prevalence was estimated based on sampling weight. Dash lines represented the overall prevalence. ID: iron deficiency; IDA: iron deficiency anaemia; MNDs: micronutrient deficiencies.

Unsurprisingly, stark differences in the nutritional status and prevalence of MNDs existed between adolescents from rural areas in comparison to urban areas (**Figure S1-3**). The prevalence of overweight was much higher in adolescents from urban areas (33·7 versus 23·6% in rural areas; **Figure S1**), while stunting prevalence was much higher in females from rural areas (17·4 versus 8·3%; **Figure S1**). The prevalence of having any MNDs was higher in all age groups from rural areas compared to those from urban areas (**Figures S2 and S3**).

### Associations between micronutrient deficiencies and demographic, socioeconomic indicators and growth indices

Logistic regressions were performed to examine the association between MNDs (individually and combined) and age, ethnicity, area of residence, wealth index and growth indices (**Table 3**). The associations between MNDs and growth indices were further estimated while adjusting for other variables that showed significant association with MNDs and variables reported to be risk factors of MNDs in previous studies (**Table 4**).

**Table 3.** Unadjusted binominal and multi-nominal logistic regression between micronutrient status and body weight status among female adolescents aged 10 to 18 years old living in Vietnam (n=1471).

|  | Individual MNDs<sup>a</sup> |  |  |  |  | Any MNDs<sup>b</sup> |  |
| --- | --- | --- | --- | --- | --- | --- | --- |
|  | Anaemia | ID | IDA | Low serum zinc | Low serum retinol | 1 MND | ≥2MNDs |
|  | COR [95% CI] | COR [95% CI] | COR [95% CI] | COR [95% CI] | COR [95% CI] | CRR [95% CI] | CRR [95% CI] |
| Age group (ref:10-12 y) | 1 | 1 | 1 | 1 | 1 | 1 | 1 |
| 13-15 y | 1.78 [0.94, 3.38] | **2.93 [1.68, 5.12]\*\*** | 2.72 [0.92, 8.06] | 0.93 [0.73, 1.17] | 1.00 [0.25, 4.04] | 1.05 [0.77, 1.43] | 2.03 [0.62, 6.69] |
| 16-18 y | **2.24 [1.40, 3.60]\*\*** | **4.73 [2.20, 10.17]\*\*** | **4.55 [1.94, 10.68]\*\*** | 1.39 [0.88, 2.20] | 0.54 [0.04, 6.93] | 1.38 [0.57, 3.31] | **4.55 [1.27, 16.28]\*** |
| Ethnicity (ref: Kinh) | 1 | 1 | 1 | 1 | 1 | 1 | 1 |
| Minorities | 1.81 [0.92, 3.54] | 0.74 [0.32, 1.71] | 1.09 [0.44, 2.7] | **1.63 [1.08, 2.47]\*** | 1.47 [0.56, 3.81] | 1.55 [1.00, 2.43] | 1.32 [0.49, 3.57] |
| Area of residence (ref: Urban) | 1 | 1 | 1 | 1 | 1 | 1 | 1 |
| rural | 1.34 [0.61, 2.96] | 1.32 [0.74, 2.35] | 1.46 [0.66, 3.23] | 1.39 [0.74, 2.61] | 1.64 [0.40, 6.69] | 1.22 [0.69, 2.15] | 1.69 [0.49, 5.81] |
| Wealth index (ref: Lowest) | 1 | 1 | 1 | 1 | 1 | 1 | 1 |
| Second | 1.03 [0.63, 1.69] | 1.59 [0.86, 2.94] | 1.33 [0.59, 3.01] | 0.95 [0.46, 1.95] | 1.72 [0.38, 7.78] | 1.41 [0.68, 2.95] | 1.77 [0.62, 5.03] |
| Third | **0.42 [0.23, 0.77]\*** | 0.77 [0.34, 1.74] | 0.62 [0.18, 2.09] | **0.48 [0.3, 0.77]\*\*** | 1.75 [0.30, 10.18] | 0.73 [0.43, 1.22] | 0.23 [0.04, 1.29] |
| Fourth | 0.52 [0.26, 1.04] | 1.32 [0.61, 2.86] | 0.95 [0.40, 2.22] | 0.89 [0.57, 1.38] | 0.38 [0.06, 2.42] | 1.42 [0.86, 2.33] | 0.51 [0.21, 1.24] |
| Highest | **0.29 [0.14, 0.61]\*\*** | **0.26 [0.11, 0.63]\*** | **0.42 [0.22, 0.81]\*** | 0.59 [0.27, 1.29] | 0.72 [0.08, 6.18] | 0.84 [0.43, 1.65] | 0.31 [0.07, 1.30] |
| Inflammation (ref: no inflammation) | 1 | 1 | 1 | 1 | 1 | 1 | 1 |
| Chronic or acute inflammation | **2.03 [1.03, 4.01]\*** | **4.11 [2.18, 7.76]\*\*** | 1.50 [0.50, 4.53] | 1.63 [0.86, 3.07] | **7.86 [1.61, 38.30]\*** | 1.42 [0.60, 3.36] | **11.44 [3.56, 36.79]\*\*** |
| Growth indices | 1 | 1 | 1 | 1 | 1 | 1 | 1 |
| Stunted (ref: non-stunted) | 1.81 [0.93, 3.51] | 1.17 [0.55, 2.49] | 1.85 [0.94, 3.65] | 1.51 [0.90, 2.53] | 3.96 [0.97, 16.18] | **1.97 [1.21, 3.20]\*\*** | **3.16 [1.09, 9.19]\*** |
| Overweight (ref: normal weight) | 0.68 [0.40, 1.18] | 1.14 [0.57, 2.27] | 0.96 [0.42, 2.20] | **0.76 [0.59, 0.99]\*** | 2.04 [0.81, 5.12] | 0.68 [0.41, 1.12] | 1.00 [0.44, 2.29] |
| Thinness (ref: normal weight) | 1.44 [0.74, 2.8] | **0.19 [0.04, 0.83]\*** | 0.24 [0.03, 2.11] | 1.04 [0.56, 1.95] | **9.15 [2.28, 36.68]\*\*** | 1.18 [0.56, 2.51] | 0.86 [0.13, 5.59] |

<sup>a</sup>Anaemia was defined as haemoglobin <115 g/L and <120 g/L for 10-11 years old and 12-18 years old, respectively. ID was defined as serum ferritin<15µg/L or 70µg/L for apparently healthy individuals and individuals with infection or inflammation [CRP >5mg/L and/or AGP >1mg/L], respectively; IDA was defined as the co-existence of anaemia and ID. Low serum zinc was defined as serum zinc under different conditions according to IZiNCG; Low serum retinol were defined as serum retinol concentration <0.70µmol/L.
<sup>b</sup>Any MNDs referred to having any of the following micronutrient deficiencies: iron deficiency, low serum zinc or retinol.
CI: confidence interval; COR: crude odds ratio; CRR: crude relative risk; ID: iron deficiency; IDA: iron deficiency anaemia; MND: micronutrient deficiencies.
\*:p<0.05; \*\*:p<0.01.

**Table 4.** Adjusted binominal and multi-nominal logistic regressions between micronutrient status and growth indices among female adolescents aged between 10 to 18 years old (n=1471).

|  | Individual MNDs<sup>1</sup> |  |  |  |  | Any MNDs<sup>2</sup> |  |
| --- | --- | --- | --- | --- | --- | --- | --- |
|  | Anaemia | ID | IDA | Low serum zinc | Low serum retinol | 1 MND | ≥2 MNDs |
|  | AOR [95% CI] | AOR [95% CI] | AOR [95% CI] | AOR [95% CI] | AOR [95% CI] | ARR [95% CI] | ARR [95% CI] |
| Non-stunted | 1 | 1 | 1 | 1 | 1 | 1 | 1 |
| Stunted | 1.42 [0.80, 2.51] | 1.03 [0.50, 2.11] | 1.51 [0.59, 3.91] | 1.54 [0.91, 2.61] | **8.92 [2.26, 35.15]\*\*** | **1.88 [1.13, 3.12]\*** | 2.30 [0.60, 8.86] |
| Normal weight | 1 | 1 | 1 | 1 | 1 | 1 | 1 |
| Overweight | 0.68 [0.36, 1.31] | 1.16 [0.55, 2.44] | 1.23 [0.46, 3.27] | 0.79 [0.58, 1.06] | 3.57 [0.78, 16.38] | 0.68 [0.40, 1.13] | 0.98 [0.42, 2.27] |
| Thinness | 1.71 [0.97, 2.99] | **0.22 [0.05, 0.99]\*** | 0.35 [0.04, 3.45] | 1.15 [0.58, 2.28] | **12.25 [3.47, 43.33]\*\*** | 1.26 [0.56, 2.83] | 1.1 [0.17, 7.13] |

All analyses were adjusted for confounders including age, ethnicity, area, wealth index, and inflammation.
<sup>1</sup>Anaemia was defined as haemoglobin <115 g/L and <120 g/L for 10-11 years old and 12-18 years old, respectively. ID was defined as serum ferritin <15µg/L or 70µg/L for apparently healthy individuals and individuals with infection or inflammation [CRP >5mg/L and/or AGP >1mg/L], respectively; IDA was defined as the co-existence of anaemia and ID. Low serum zinc was defined as serum zinc under different conditions according to IZiNCG; Low serum retinol were defined as serum retinol concentration <0.70µmol/L.
<sup>2</sup>Any MNDs referred to having any of the following micronutrient deficiencies: iron deficiency, low serum zinc or retinol.
AOR: adjusted odds ratio; ARR: adjusted relative risk; CI: confidence interval; ID: iron deficiency; IDA: iron deficiency anaemia; MND: micronutrient deficiencies.
\*:p<0.05; \*\*:p<0.01.

Our findings from bivariate logistic regression showed that while female adolescents aged 13-15 years old had increased odds of ID (COR = 2·93 [1·68, 5·12], p<0·01), those aged 16-18 had much higher odds of having IDA (COR = 4·55 [1·94, 10·68], p<0·01), and more than one MND (COR = 4·55 [1·27, 16·28], p<0·01; **Table 3**). Inflammation status was associated with increased odds of anaemia (COR = 2·03 [1·03, 4·01], p<0·05), ID (COR = 4·11 [2·18, 7·76], p<0·01), and low serum retinol (COR = 7·86 [1·61, 38·30], p<0·05), but had no impact on odds of low serum zinc (**Table 3**). Ethnic minorities had increased odds of low serum zinc (COR = 1·63 [1·08, 2·47], p<0·05); whereas female adolescents from the wealthiest quintile were significantly protected against IDA (COR = 0·42 [0·22, 0·81], p<0·05). Interestingly, no significant association was found between MNDs and rural or urban areas of residence.

In the multivariate logistic regression models, after adjusting for age, ethnicity, area of residence, wealth index and inflammation; stunting and thinness were associated with increased odds of low serum retinol (stunting: AOR: 8·92 [2·26, 35·15], p<0·01; thinness: AOR: 12·25 [3·47, 43·33], p<0·01). In addition, stunting increased odds of having at least one MND (AOR: 1·88 [1·13, 3·12], p<0·05) compared to those without any MNDs (**Table 4**). However, it is important to note that the prevalence of low serum retinol among the study participants was extremely low (1·5%, n=16; Table 4), thus these results should be interpreted with caution. The findings also suggest a marginally significant protective effect of thinness against the odds of ID (AOR: 0·22 [0·05, 0·99], p=0·049). We found no significant impact of any growth indices on the odds of low serum zinc (**Table 4**). In the unadjusted bivariate logistic regression, overweight appeared protective against low serum zinc (COR: 0·76 [0·59, 0·99], p=0·04; **Table 3**). However, this association disappeared after adjusting for covariates (**Table 4**). Similarly, no significant association was found between overweight and any MNDs in the multivariate logistic regression models. The results of secondary analysis on the randomly generated subsamples (50%) were consistent with the results on the full dataset reported above.

## DISCUSSION

This is the first study to investigate the prevalence and associations between MNDs and nutritional status (i.e., both under-and overnutrition), in contempory female adolescents living in Vietnam. Our data starkly illustrate the current double burden of malnutrition in Vietnam, highlighting a high prevalence of overweight (27%) co-existing alongside undernutrition (approximately 22%; combined prevalence of stunting (14·3%) and thinness (6·9%)) among female adolescents. Low serum zinc was the most commonly observed MND, affecting 40% of participants in this cross-sectional cohort, followed by ID (9·8%). In contrast, low serum retinol was rarely observed (1·5%). Notably, stunting significantly increased the risk of having any MND (ARR: 1·88 [1·13, 3·12]), and the prevalence of stunting still predominated over overweight (26·8% versus 22%) in late adolescent females aged 16-18 years old living in Vietnam in 2020.

Between 2010 and 2020 the prevalence of overweight in children and young people (5-19 years old) living in Vietnam was estimated to have more than doubled (8·5 to 19%).^7^ Strikingly, our study suggests an even higher prevalence of overweight among female adolecents living in Vietnam between 2019 and 2020, reaching 27%. This is higher than most LMICs in the South-Eastern Asia region,^7^ and likely stems from a marked nutrition transition in Vietnam that has come secondary to a period of rapid and sustained economic growth with large increases in household wealth.^7,16,17^ Our study illustrates the scale of the double burden of malnutrition in female adolescents in Vietnam, as a comparable percentage of participants in this nationally-representative survey were underweight as overweight. This emergent public health problem has been prioritised in the National Nutrition Strategy for 2021-2030.^16^

Significant demographic differences were observed between stunted and non-stunted adolescents, this included participant’s areas of residence (urban versus rural), geographical regions, ethnicity, and wealth index. These factors are often interrelated, as in Vietnam many ethnic minorities live in rural or mountainous areas, with limited education, household income and diet choice. Minority groups are much more likely to live in moderate or severe food insecurity, and have less access to health care services including vaccinations, as well as the improved sanitation and water supply found in urban regions.^16,18^ Ethnic minorities consequently have a higher prevalence of parasitic infections and diarrhea, leading to increased risk of stunting in early childhood (under 5 years of age).^19^ Therefore, in the Vietnam Nutrition Strategy 2021-2030, ethnic minorities were seen as a critical target population to be individually evaluated for most nutrition targets.^16^ Our data showing that wealth index was higher in non-stunted adolescents aligns with previous research that linked household wealth to protection from childhood stunting in developing countries including Vietnam.^20^

Interestingly, we found no significant association between body weight status (thinness, normal, or overweight) and either wealth index, ethnicity, geographical regions, or areas of residence. That is, the results show a high prevalence of overweight among Vietnamese female adolescents irrespective of their sociodemographic factors. These data are in contrast to the Young Lives study (Vietnamese older cohort), which found the children within the top tertile of household wealth had a much higher relative risk (RR) of overweight and obesity in comparison to those in the bottom tertile, albeit with a wide 95% confidence interval (RR: 9·11 [1·07, 77·42]).^21^

In addition to negatively impacting immune function and increasing the risk of morbidity for the adolescents themselves, MNDs, in female adolescents in particular, can lead to intergenerational consequences through shaping fetal programming, development in early life, and the cardiometabolic health of the offspring in the long term.^3^ Our study highlights a high prevalence of low serum zinc levels and ID in Vietnamese female adolescents, with those from higher household wealth being protected from ID, IDA, and all-cause anaemia. The much lower prevalence of low serum retinol may be attributed to the universal high dose vitamin A supplement programs (100,000 IU) for children aged 6-36 months, which have been implemented in Vietnam since 1988.^22^ Female adolescents from minority ethnic groups were more likely to have low serum zinc, likely related to both increased prevalence of food insecurity, infections, and diarrhea, as well as consumption of more staple and starchy foods that are lower in essential micronutrients.^18^

Although with economic growth, the dietary pattern of Vietnamese has changed to include more meat, vegetables and less starchy food at a national level, stark differences in household expenditure on food groups exist between ethnic minority, rural, and poorer households.^6,18^ Additionally, although the GNS 2020 followed international guidelines for defining MNDs (e.g., adjusting ferritin thresholds based on CRP and AGP measurements for inflammation, and adjusting cut-offs for low serum zinc depending on sample type (i.e. fasted versus non-fasted) and sample collection time); nonetheless, we caution multiple challenges with MND assessment and zinc status measurement in particular exist.^23^ For example, serum zinc levels may be transiently lowered in acute infection or inflammation, which can lead to overestimation of deficiency. Conversely, haemolysis or contamination from adventitious zinc in the environment could mask low serum values.

Notably, we found that MNDs were generally more prevalent among older adolescents (16-18 years old), which may reflect increased demands for both micro-and macronutrients during growth spurts and sex maturation during puberty.^24,25^ In addition, repeated blood loss during menstruation will deplete iron stores particularly for females with limited access to iron-rich foods, or who have low iron absorption, which leads to ID with or without anaemia.^26,27^ In this study, we did not find any association between overweight and MNDs, although overweight, in particular obesity, has been shown to increase risk for ID in children and young people in meta-analysis OR=1·51 (1·20, 1·82).^23^ The possible explanation could be the extremely low prevalence of obesity (2·7% in urban and 1·3% in rural area) in our population in comparison to those included in the meta-analysis, which were from North America and European populations. Our results match those of Laillou and colleagues,^28^ who concluded overweight/obesity was not a risk factor for ID in a a large cohort of Vietnamese women (n=1526).

As with all observational studies, our cross-sectional study has limitations, most notably that it can not reveal causality. In addition, fewer participants recruited to the GNS 2020 were in late adolescence, which may limit the interpretation of the logistic regression. This age group should be a particular focus for data collection in the next GNS survey. Not least, an important limitation in examining this age group was that due to missing values we could not include puberty stage as a confounder in regression analyses. Nonetheless, our study had notable strengths. The GNS 2020 was a nationally-representative population based survey, meaning our findings can be generalised to female adolescents of the same age group across Vietnam. Another strength was the measurement of biochemical biomarkers of micronutrient status and inflammation, alongside anthropometric and socio-demographic data. This study provides unprecedented insights into the current double burden of malnutrition among female adolescents in Vietnam.

In conclusion, our data highlight that more female adolescents were living with overnutrition than undernutrition in Vietnam in 2020. However, undernutrition and low serum zinc remain persistently prevalent. Regional, residential, ethnic and wealth disparities in undernutrition and MND suggest the needs for targeted interventions to ensure equitable access for all and to meet local needs. These data should aid in prioritizing public health and nutrition education initiatives, and provides reference points for assessing the effectiveness of future interventions. Robust, timely and reliable data on the burden of MNDs is essential to inform, design and implement successful nutrition policies and programmes aimed at reducing malnutrition. Food environment and food systems approaches should be considered to stem the recent stark increase in overweight in young people living in Vietnam and other Western Pacific nations newly facing the double burden of malnutrition.

### Contributors

XT, PYT, SVS, YYG and JBM designed and conceptualized the study. YYG and JBM supervised the study. NTT and VKT were responsible for GNS 2020 data curation, investigation, and methodology. SDN and DTT performed data cleaning and final statistical analyses. NTT, VKT, SDN and DTT had full access to all the data and took responsibility for the integrity of the data. XT, PYT and SVS wrote code for formal analyses and performed statistical analyses on data subsets. XT prepared the tables and figures and original manuscript draft. JBM critically revised the manuscript and all authors reviewed the manuscript and approved the submitted final version.

### Data sharing statement

The GNS 2020 dataset is the property of the National Institute of Nutrition, Vietnam, and is not publicy available. This secondary analysis was conducted under a partnership agreement between the University of Leeds and NIN. Requests to access the datasets should be directed to.

### Ethics statement

Ethical approval was not required for the secondary data analysis in this study. The GNS 2020 was reviewed and approved by the Ethical Committee of the National Institute of Nutrition, Ministry of Health, Vietnam. All methods were conducted in accordance with the guidelines laid down in the Declaration of Helsinki and written informed consent was obtained from each participant prior to data collection by NIN.

## Acknowledgments

This research was funded by the UK Biotechnology and Biological Sciences Research Council (BBSRC) Global Challenges Research Fund (B/T008989/10). The funders had no involvement in study design, data analysis or interpretation or decision to submit the paper for publication. The authors extend their gratitude to extended colleagues in the National Institute of Nutrition, Ministry of Health, Vietnam, for their roles in the GNS 2020, and huge thank you to the participants in that survey.

