## Supplementary material for "Micronutrient deficiencies and the double burden of malnutrition in Vietnamese female adolescents: a national cross-sectional study in 2020": Table S1; Figure S1-3

**Table S1 Number and percentage missed observations in dataset.**

| | Variable | Number of observations | Number of missed observations | Percentage of missed observations |
|---|---|---|---|---|
| **Age** | age in year | 1471 | 0 | 0.0 |
| **Anthropometry** | body weight | 1471 | 0 | 0.0 |
| | body height | 1253 | 218 | 14.8 |
| **MNDs & inflammation** | anemia | 1396 | 75 | 5.1 |
| | ID | 1296 | 175 | 11.9 |
| | IDA | 1396 | 75 | 5.1 |
| | low serum zinc | 1389 | 82 | 5.6 |
| | low serum retinol | 1133 | 338 | 23.0 |
| | number of MNDs | 868 | 603 | 41.0 |
| | inflammation status | 1297 | 174 | 11.8 |
| **Socioeconomic** | ecological region | 1471 | 0 | 0.0 |
| | residence of living | 1471 | 0 | 0.0 |
| | ethnicity | 1471 | 0 | 0.0 |
| | wealth index | 1471 | 0 | 0.0 |
| **Others** | puberty | 212 | 1259 | 85.6 |

ID: iron deficiency; IDA: iron deficiency anaemia.

**FIGURE S1**

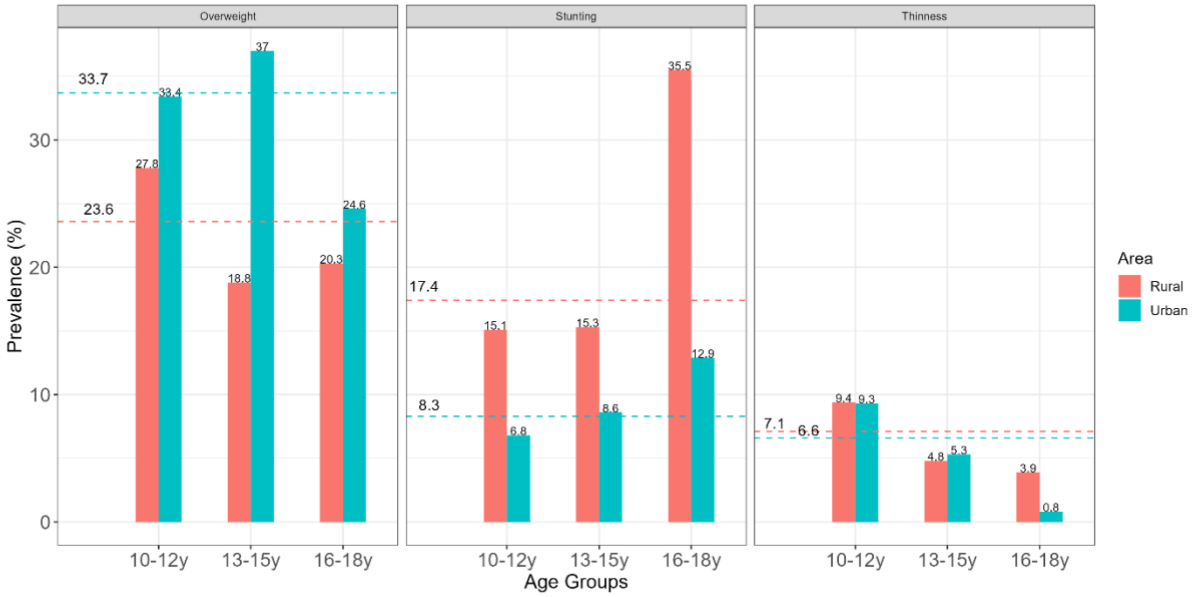

**Figure S1  Prevalence of overweight, stunting and thinness in rural and urban areas.**
Overweight: BMI-for-age z score (BAZ) > +1 SD, stunting: height-for-age z score (HAZ)< -1 SD, thinness: BAZ< -2 SD. Prevalence was estimated based on sampling weights. Bars represent the prevalence in each age groups. Dashed lines illustrate the overall prevalence.

**FIGURE S2**

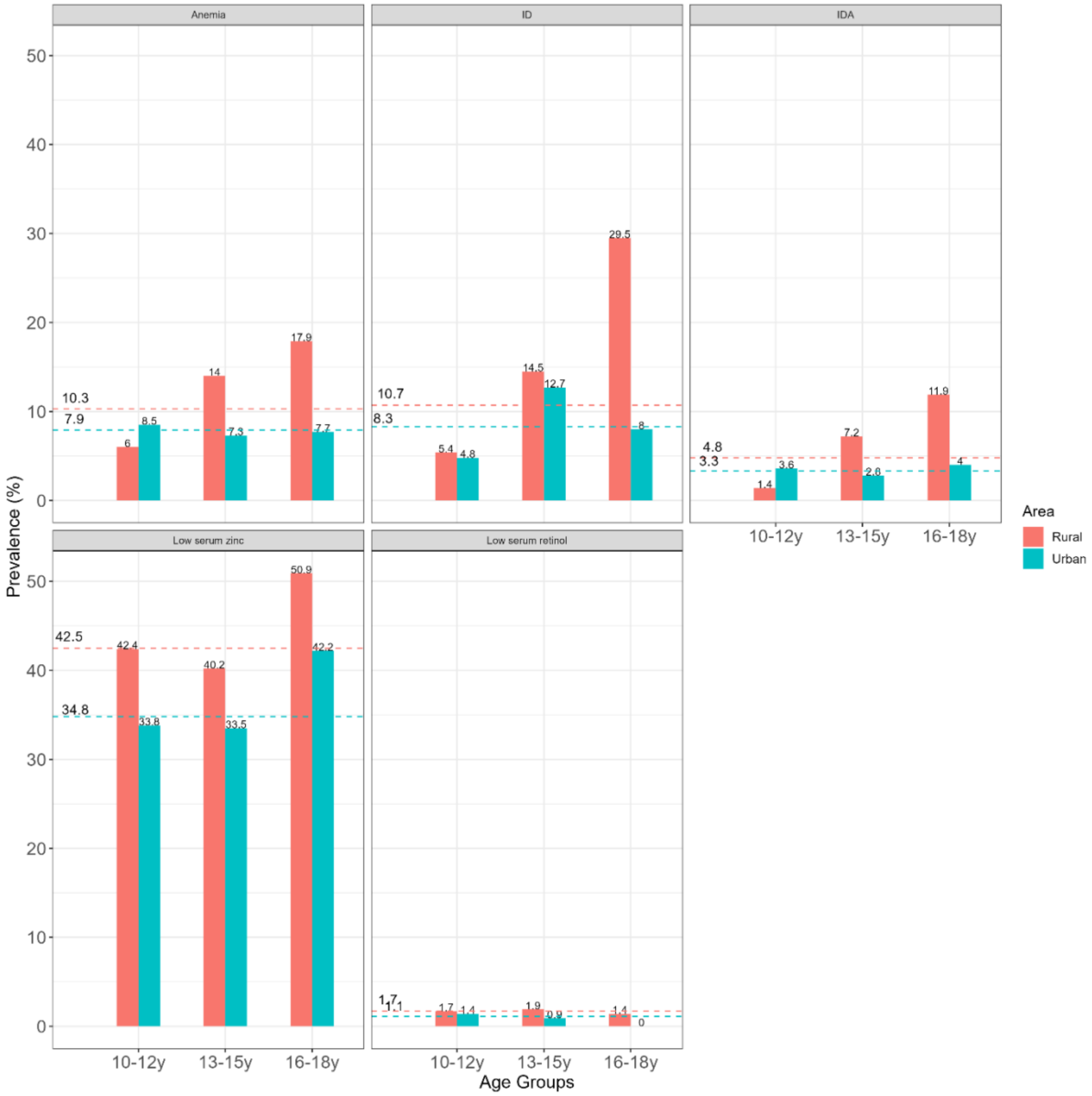

**Figure S2 Prevalence of micronutrient deficiencies in rural and urban areas.** ID: iron deficiency, IDA: iron deficiency anaemia.

**FIGURE S3**

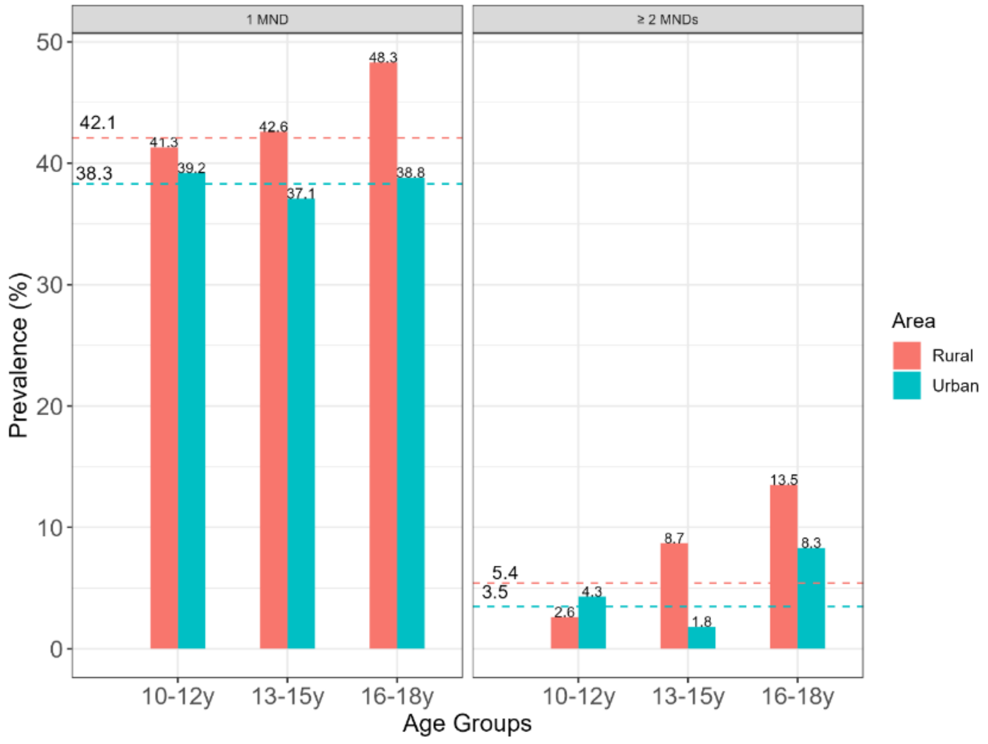

**Figure S3 Prevalence of any MNDs in rural and urban areas.** MNDs included any of: iron deficiency, low serum zinc and low serum retinol.
